# 3D Deep Neural Network Segmentation of Intracerebral Hemorrhage: Development and Validation for Clinical Trials

**DOI:** 10.1101/2020.03.05.20031823

**Authors:** Matthew F. Sharrock, W. Andrew Mould, Hasan Ali, Meghan Hildreth, Issam A. Awad, Daniel F. Hanley, John Muschelli

## Abstract

Intracranial hemorrhage (ICH) occurs when a blood vessel ruptures in the brain. This leads to significant morbidity and mortality, the likelihood of which is predicated on the size of the bleeding event. X-ray computed tomography (CT) scans allow clinicians and researchers to qualitatively and quantitatively diagnose hemorrhagic stroke, guide interventions and determine inclusion criteria of patients in clinical trials. There is no currently available open source, validated tool to quickly segment hemorrhage. Using an automated pipeline and 2D and 3D deep neural networks, we show that we can quickly and accurately estimate ICH volume with high agreement with time-consuming manual segmentation. The training and validation datasets include significant heterogeneity in terms of pathology, such as the presence of intraventricular (IVH) or subdural hemorrhages (SDH) as well as variable image acquisition parameters. We show that deep neural networks trained with an appropriate anatomic context in the network receptive field, can effectively perform ICH segmentation, but those without enough context will overestimate hemorrhage along the skull and around calcifications in the ventricular system. We trained with all data from a multi-center phase II study (n = 112) achieving a best mean and median Dice coefficient of 0.914 and 0.919, a volume correlation of 0.979 and an average volume difference of 1.7ml and root mean squared error of 4.7ml in 500 out-of-sample scans from the corresponding multi-center phase III study. 3D networks with appropriate anatomic context outperformed both 2D and random forest models. Our results suggest that deep neural network models, when carefully developed can be incorporated into the workflow of an ICH clinical trial series to quickly and accurately segment ICH, estimate total hemorrhage volume and minimize segmentation failures. The model, weights and scripts for deployment are located at https://github.com/msharrock/deepbleed. This is the first publicly available neural network model for segmentation of ICH, the only model evaluated with the presence of both IVH and SDH and the only model validated in the workflow of a series of clinical trials.

## Introduction

Intracranial hemorrhage (ICH), also known as hemorrhagic stroke, accounts for 15% of all strokes, but has a significantly higher morbidity and mortality, in part, because of a lack of effective early intervention strategies. ICH occurs when blood spontaneously breaches the confines of the cranial vessels and enters into the surrounding tissue. This extravasation leads to the formation of a hematoma that may produce complications such as inflammation, edema and mass effect leading to secondary brain injury(Hanley 2009). Afterwards, ICH may extend into intraventricular areas, denoted as intraventricular hemorrhage (IVH) as it extends to the cerebrospinal fluid filled ventricular system during the natural course of the disease. A small number of patients with primary ICH may also develop a subdural hematoma (SDH) as a consequence of the initial event.

Delineating the presence of hemorrhage and calculating its volume using geometric estimation equations is a routinely performed task, albeit prone to manual error, in the care of ICH patients(Qureshi et al. 2009), as it is a consistent predictor of mortality and long-term functional outcome(Broderick et al. 1993; Hemphill et al. 2001; Tuhrim et al. 1999; LoPresti et al. 2014; Anderson et al. 2008). X-ray computed tomography (CT) is utilized as the standard imaging modality for ICH, as the acquisition of images is fast and the hematoma shows strong contrast with healthy brain tissue.

Expedient decision making is required and simplified linear metrics are often utilized to make an estimate of volume(Broderick et al. 1993; Kothari et al. 1996) and have shown to be reliable with smaller, deep hemorrhages that fit the underlying assumption of a spherical or ellipsoid shape, but less so with large, complex hemorrhages(Webb et al. 2015). Despite substantial limitations, estimation is routinely used to inform decisions about surgical intervention, aid discussions of prognosis and in clinical trials, serve as an inclusion criteria, an adaptive randomization factor, primary(Anderson et al. 2008; Mayer et al. 2005; Hussein et al. 2013) or secondary outcome measure(Anderson et al. 2008; Garg et al. 2012; Morgan et al. 2008). The Minimally Invasive Surgery Plus Alteplase for ICH Evacuation (MISTIE) clinical trials utilized highly precise core lab segmentations to produce ICH volumes. They were designed to test the safety and efficacy of an experimental therapy using minimally invasive surgical techniques to remove ICH hematoma volume. To date, the most recent phase III trial found that when patients had the volume of hematoma reduced by more than 70% or to less than 15mL of residual volume, they had improved functional outcome(Hanley et al. 2019).

Deep neural networks (DNNs) have recently improved the quality of segmentation of pathology on medical images including ischemic stroke on CT perfusion studies(Maier et al. 2017) and brain tumors on MRI(Maier et al. 2017). DNNs are trained to perform computer vision tasks by encoding the information in the scan, decoding it automatically, and then utilizing the difference between the decoded estimate and human labelled data to learn the task(LeCun et al. 2015). Several alternative methods have been presented as automated methods for ICH segmentation from CT scans including fuzzy clustering, level-set thresholds and decision-tree analysis(Prakash et al. 2012; Gillebert et al. 2014; Muschelli et al. 2017; Loncaric et al. 1995). To our knowledge, only Gillebert, Humphreys, and Mantini(Gillebert et al. 2014) and our previous work in Muschelli et al.(Muschelli et al. 2017) have made publicly available software to implement the segmentation (https://github.com/muschellij2/ichseg). Previous studies have trained deep networks to perform the task of ICH segmentation on CT(Ironside et al. 2019; Chang et al. 2018; Islam et al. 2018). However, they were trained on single institutional datasets and primarily utilized 2D networks. We hypothesized that 3D segmentation may improve results and we compare 3D models with a 2D network and our previously published random forest model.

In this study we utilized gold standard, human labelled data directly from the phase II and phase III multi-center MISTIE clinical trials. We hypothesized that we could successfully train neural network models on the earlier, smaller phase II trial and then validate on the phase III trial. This process mimics the workflow used in clinical trials, where phase II studies primarily test safety in a smaller population and phase III studies test efficacy in a larger population. In order to optimize the network for clinical trial applications and improve external validity, we combined standard neuroimaging preprocessing techniques such as brain extraction, registration and resampling as well as DNN techniques including estimates of the anatomic information presented within the networks receptive field, data augmentation and regularization in order to optimize the models for clinical trial deployment.

Our final model is a 3D neural network we call DeepBleed and we make the model and its trained weights available for external use and testing (https://github.com/msharrock/deepbleed). This is the first such deep network for ICH segmentation made publicly available and the only one with either inclusion of IVH and SDH or trained and validated on multi-center data. The model performs at the level of the current state of the art while accommodating the diversity of data inherent in clinical trials.

## Methods

### Data

Our data was derived from the phase II and phase III MISTIE multi-center randomized controlled clinical trials. Inclusion criteria were the same for both trials and included: 18 to 80 years of age and spontaneous supratentorial intracerebral hemorrhage(Mould et al. 2013; Hanley et al. 2019). The CT data was collected as part of the Johns Hopkins Medicine IRB-approved MISTIE research studies with written consent from participants.

For training, we used non-contrast CT images from patients enrolled in the MISTIE II clinical trial(Hanley et al. 2016). We analyzed 112 scans (N=112) taken prior to randomization and treatment, corresponding to the first scan acquired post-hemorrhage. We randomly selected 100 scans for model training and12 (11%) scans for testing.

For validation of the model, we used data from the MISTIE III trial(Hanley et al. 2019). We analyzed 500 scans (N = 500), again taken prior to randomization and treatment, corresponding to the first non-contrast CT scan acquired post-stroke. No data from this validation set was used in training or estimating the model and should represent how this model performs in general on multi-center data acquired during a clinical trial. The same processing steps were performed for the MISTIE II and III data. There were 77 (69%) males in MISTIE II and 304 (61%) in MISTIE III; the mean (SD) age was 60.7 (11.2) years in MISTIE II and 61.2 (12.3) years in MISTIE III. Additional data regarding the demographics of the underlying data is in the supplemental materials.

As each data set was collected from multiple sites, a number of different scanning protocols were used. The images were all non-contrast CT images with a soft-tissue convolution kernel, commonly sampled with a large thickness (e.g. 5mm) in the axial plane. The patients were from 26 different scanning sites in MISTIE II and 78 different sites (from 9 different countries) in MISTIE III, with 17 sites shared between the 2 studies.

In MISTIE II, the different scanner types were: GE (N=46), SIEMENS (N=38), Philips (N=20), and TOSHIBA (N=8). There were 88 scans (79%) with gantry tilt and 14 scans (12%) with variable slice thickness. The scans used tube voltages of either 120, 130, or 140 kVp, had within-plane pixel resolution ranging from 0.37-0.58mm and a slice thickness ranging from 0.625 to 6 mm.

In MISTIE III, the different scanner types were: GE (N=185), SIEMENS (N=166), Philips (N=72), TOSHIBA (N=71), NeuroLogica (N=5), and Hitachi (N=1). There were 283 scans (57%) with gantry tilt and 32 scans (7%) with variable slice thickness. The scans used tube voltages of either 100, 110, 120, 130, 135, or 140 kVp, had within-plane pixel resolution ranging from 0.29-0.67mm and a slice thickness ranging from 0.41 to 8 mm.

Ground truth was estimated by manual segmentation on CT scans using the OsiriX imaging software by a core laboratory staffed with expert readers (OsiriX v. 4.1, Pixmeo; Geneva, Switzerland). See Muschelli et al. (2017) for additional details. In MISTIE II, the ground truth mean (SD) hemorrhage volume was 43.2 (23.9) mL (range 5-141mL); in MISTIE III, it was 47.4 (21.4) mL (range 1-145mL).

### Image Processing

The overall processing pipeline is outlined in Figure 1. CT images and binary hemorrhage masks were exported from OsiriX to DICOM (Digital Imaging and Communications in Medicine) format and were converted to NIfTI (Neuroimaging Informatics Technology Initiative) format using dcm2niix (Li et al. 2016). Images were constrained to values −1024 and 3071Hounsfield units (HU) to remove potential image rescaling errors and artifacts.

**Figure 1:**
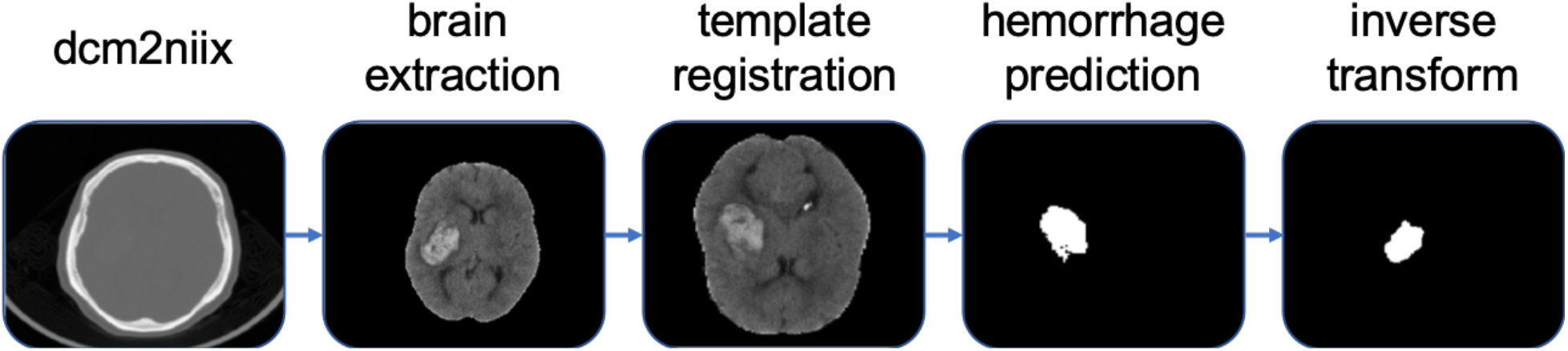
Processing pipeline for the 3D DNN models in our study. Non-contrast CT DICOM series are processed by the dcm2niix package where gantry tilt and unequal slices are corrected and normalized and the image is converted into the NIfTI format. Then using the ANTs software package, the scans are registered to a 1.5mmx1.5mmx1.5mm template. The BET tool from the FSL software package is then used to perform brain extraction. The data is then fed into the VNet and an inference is generated in the template space. Finally the prediction undergoes the inverse transform from the prior registration into the native space of the original NIfTI scan. The 2D 512 network only included conversion to NIfTI format with dcm2niix before being fed into the 2D VNet.

Image analysis was done in the R statistical software (R Core Team 2015) and Python (Python software Foundation, version 3.6). In R, the ichseg package was used (https://github.com/muschellij2/ichseg), using the fslr package(Muschelli et al. 2015a) to call functions from the FSL(Jenkinson et al. 2012) neuroimaging software (version 5.0.10) and the ANTsR package to call functions from the ANTs (Advanced Normalization Tools) neuroimaging software(Avants et al. 2011). Statistical analyses and figures were created in R. Brains were extracted to remove skull, eyes, facial and nasal features, extracranial skin, and non-human elements of the image captured by the CT scanner, such as the gurney, pillows, or medical devices. Removal of these elements was performed using the brain extraction tool (BET)(Smith 2002), a function of FSL, using a previously published validated CT-specific brain extraction protocol(Muschelli et al. 2015b). For the 3D neural networks, images were registered to a 1.5×1.5×1.5mm CT template(Rorden et al. 2012) using a rigid-body registration and a nearest neighbor interpolation. The dimensions of template are 128×128×128 voxels. This was to allow for z-axis normalization without under sampling scans with higher resolution. For the 2D neural networks, the scans were segmented in their native space. All scans were acquired in a 512×512 field. Scans with gantry tilt, after correction by dcm2niix, had x and y dimensions that were not 512×512 and were resampled to a 512×512 space before running the model.

Areas outside the brain mask were set to 0 and no further normalization was applied. We felt this was justified as HU measurements are calibrated with pure water set to a value of 0 for all scanners and the range of values that encompass both normal brain tissue (∼20 - 60HU) and ICH (∼40 - 100HU) are in similar regions of the HU scale with the same order of magnitude. Performing brain extraction also helps to normalize the image intensities as air (- 1000 HU) and bone (∼ 1000 HU) are set to 0.

### Models

In this study, we fit fully convolutional neural networks (CNNs) with Tensorflow(Abadi 2016) (version 2.0.0) in Python (version 3.6). We fit models using a modified VNet framework(Milletari et al. 2016). The main modification to the network was the addition of dropout into the deepest layers to provide additional regularization and a binary sigmoid output layer. For the 2D network, we dropped the 3rd dimension in each convolutional filter.

We fit one 2D network and three 3D networks. Each network was categorized by a patch size indicating how much of the original image is fed into the network with each pass. In the 2D network, we used the 512^2^ slices individually, and call this model the ***512 network***.

For feeding into the 3D networks, the total input patch size was different for each network in order to vary the amount of contextual anatomy presented within the networks receptive field (RF). The RF of a neural network is the number of input pixels that influence the value of one output pixel. The VNet architecture was designed such that the RF of the network spans the entire input(Milletari et al. 2016). Each convolutional block on the left side of the network doubles the RF until the entire input image influences the output. The portion of the RF that has the strongest impact on output is termed the effective receptive field (ERF), is gaussian distributed from the center pixel of the RF and is not influenced by the addition of dropout layers(Luo et al. 2016).

We hypothesized that in order for the network to learn normal CT findings such as calcifications in the ventricular system or averaging with the skull at the cortical surface we would need to provide the anatomic context to distinguish between the cortical surface and deeper structures. Assuming a brain with a width of 150mm and a normal ventricular width ratio of less than 0.30, we would need a sample of width > 50mm to reliably sample either the brain surface or the ventricular system(Jaraj et al. 2017). We trained three 3D networks with different sampling sizes. The network that processed all the resampled data at once (128^3^) we named the ***128 network***, one utilizing input blocks of 64^3^ we named the ***64 network*** and the one utilizing smaller 32^3^ blocks we named the ***32 network***. The real world spatial lengths of the networks are 196mm, 96mm and 48mm respectively.

While training, to reduce overfitting on the training data both data augmentation and dropout regularization were utilized. Dropout was utilized in the deepest layers of the network. Our training strategy included the use of small initial learning rates and the adaptive moment estimation (Adam) stochastic optimizer as otherwise the networks converged quickly on the training data in the first 100 epochs, leading to poor performance on the testing data. In order to prevent overfitting we utilized augmentation with random left-right flipping and random elastic deformation without z-axis deformations given the relatively low z-axis resolution in our dataset. We used mini-batches to train the networks in all of the 2D and 3D implementations. For the 512 network, due to the class imbalance in the dataset, weighted sampling was used where the likelihood of sampling a slice was determined by the proportion of pixels inside the label and the loss function utilized was cross entropy.

For the 3D networks, the Dice loss function as implemented in the original VNet paper was used(Milletari et al. 2016). We trained all networks for a minimum of 200 epochs and thereafter, a decision to stop training was made once the moving average loss on the testing data did not improve over the previous 10 epochs. The networks were trained using CPU multithreading and two Nvidia Tesla P100 GPUs. The training time was approximately 16 hours to reach 200 epochs for all CNN models.

### Analysis

Each network produces a binary segmentation of ICH and IVH if present, for each scan. In the case of the 512 network, the 2D slices are processed and then stacked into a 3D prediction. For the 64 and 32 networks, an overlap with averaging was employed to eliminate the effect of edge cases and reassembled into a prediction with a 128^3^ volume. We also segmented the images using PItcHPERFeCT(Muschelli et al. 2017) to compare results without any retraining of the model. All of the 3D model predictions were transformed back into the native space using the inverse registration transformation. All comparisons and performance measures calculated in native space.

To estimate the performance of the model, we compared the automated segmentation and manual segmentation from the MISTIE clinical trial. To estimate volumetric overlap, we calculated the Dice Coefficient (DSC)(Dice 1945), for absolute volume differences, the mean-squared error (MSE), for relative volume differences, the correlation of automated versus manual volumes. We calculate these measures for both the training and validation data. The validation results provide a measure of external validity; comparing these results to the training results provide a measure of overfitting.

## Results

Of the 506 patients were enrolled in the MISTIE III trial, 6 patients did not have their CT scan immediately available at the time of this study. We therefore analyzed 500 patient scans. 276 (55%) had ICH only, 205 (41%) had IVH, 16 (3%) had SDH, and 4 (1%) had both IVH and SDH. No scans were excluded due to image quality. We present the results for the MISTIE III data in Table 1 and the overall distribution of DSC in Figure 2. We note the 64 and 128 networks perform similarly and the best overall with a mean (SD) DSC of 0.910 (0.04) and 0.911 (0.04) for each model respectively. The 512 network had a lower performance of 0.899. Using the original PItcHPERFeCT model on the MISTIE III validation data resulted in a mean (SD) Dice of 0.881 (0.08).

**Table 1:** Dice Coefficient (DSC) statistics for all four models on the MISTIE III dataset. We note the 3D 64 and 128 models perform best and similar to each other. We do note, however, that the 2D 512 model has a higher minimum DSC compared to other models. We define a failure as a DSC < 0.5. All model results are from the 500 patient scans in the validation set.

| Dice Coefficient Statistics |  |  |  |  |  |  |
| --- | --- | --- | --- | --- | --- | --- |
| Model | Mean | SD | Median | Min | Max | Percent Failed |
| **2D VNET 512** | 0.899 | 0.047 | 0.910 | 0.490 | 0.963 | 0.0% |
| **3D VNET 128** | 0.910 | 0.039 | **0.919** | **0.582** | 0.955 | 0.0% |
| **3D VNET 64** | **0.911** | 0.040 | **0.919** | 0.544 | 0.958 | 0.0% |
| **3D VNET 32** | 0.788 | 0.101 | 0.817 | 0.165 | 0.936 | 2.0% |
| **PltchPERFeCT** | 0.881 | 0.078 | 0.902 | 0.000 | **0.966** | 1.0% |

**Figure 2:**
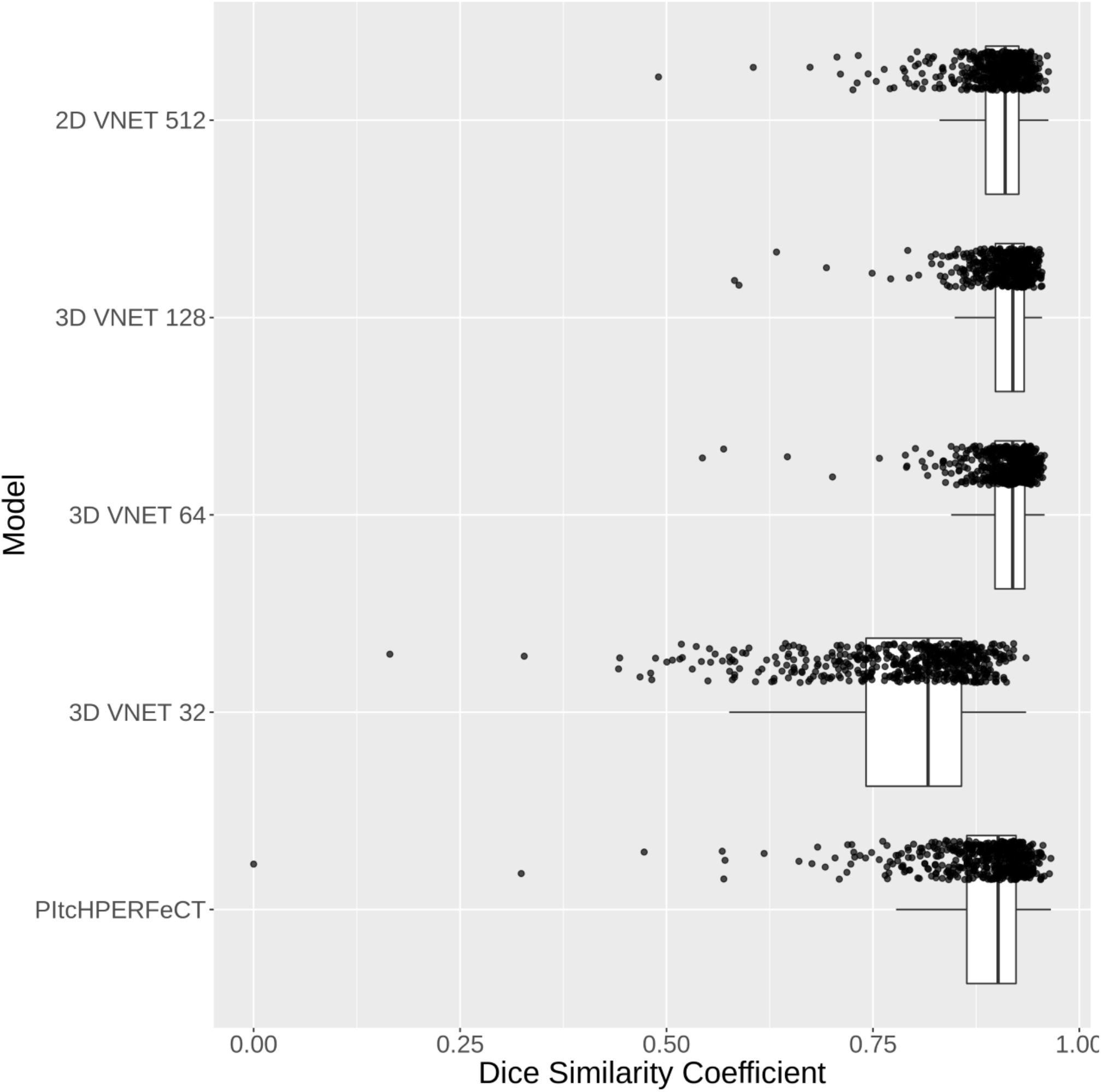
Distribution of Dice Coefficients for Each Model. Here we present the Dice Coefficient/Similarity Index (DSC) for all patients in the 501 MISTIE III validation set. We see that the 32 network performs poorly, the 2D (512) network and the 3D 128 and 64 networks perform well and comparably, and the PItcHPERFeCT performs reasonably well with more variability and lower median performance compared to all networks other than the 32 network.

As we believe the blood location and type may affect performance, we will report both the overall results for each method along with the breakdown for each hemorrhage combination subtype. We defined a segmentation failure as a DSC < 0.5. No failures occurred in the MISTIE III data for the 2D and 3D networks except for the 32 network with 2 failed scans. The overall mean and standard deviation of the DSC for each model and hemorrhage subgroup is reported in Table 2. Overall, these results indicate good performance compared to manual readers for estimating intracranial blood from CT.

**Table 2:** Dice Coefficient mean and standard deviation for hemorrhage subgroups in the MISTIE III dataset. The model outputs are compared to the gold standard, hand segmentation.

| Mean (SD) Dice Coefficient |  |  |  |  |
| --- | --- | --- | --- | --- |
| Model | ICH only | ICH with IVH | ICH with SDH | ICH with IVH and SDH |
| **2D VNET 512** | 0.9 (0.052) | 0.902 (0.035) | 0.862 (0.057) | 0.889 (0.038) |
| **3D VNET 128** | **0.914 (0.041)** | 0.908 (0.035) | **0.879 (0.049)** | 0.892 (0.043) |
| **3D VNET 32** | 0.781 (0.113) | 0.802 (0.082) | 0.711 (0.078) | 0.775 (0.092) |
| **3D VNET 64** | 0.913 (0.045) | **0.911 (0.027)** | 0.869 (0.076) | **0.898 (0.035)** |

### Volume Comparison

Although the DSC gives an indication of overlap between the predicted segmentation and ground truth, total volume is a comparator that is also a predictor of functional outcome. Moreover, the volume of ICH is an inclusion criterion for many hemorrhagic stroke studies. DSC can vary wildly for ICH that is small (< 5mL), though the absolute volume difference can be small.

We present the results for each model in Table 3. Overall, the correlation between the automatic volume and the manual volume is high for all networks except for 32 network and PItcHPERFeCT. We can see these high correlations reflected in Figure 5, which compares the volumes in a scatterplot. The black X=Y line represents perfect agreement, whereas the blue line represents a linear fit and the red line represents a smoothed fit to the data. Though the correlation is not 1, we note that the overall shape of the fit for each model is linear (blue and red line agreement), indicating that using the automated volume in linear models should provide similar results, on average. The root mean squared error (RMSE) in Table 3 represents the average volume difference between the 2 measurements, which is around 5-8mL. This agreement it close to 5mL, similar to the the inter-reader variability from estimating ICH manually(Kothari et al. 1996; Huttner et al. 2006; Hussein et al. 2013), although larger differences have been observed between clinical sites and the trial center(Webb et al. 2015). Using the threshold of 5mL, we see that above 72% of the patient scans fall within this range for the **64, 128**, and **512** networks.

The average volume difference also indicates that the automated measurements underestimate the true volume by 2-4mL (Table 3). We investigated where this underestimation occurs in a Bland-Altman plot in Figure 4, which displays the average of the automated and manual volume compared to its difference (bias). The blue line represents perfect agreement, the red lines represent the 5mL limits, and the pink line represents a smoothed fit to the data. Divergence of this pink line from the horizontal represents that the bias in the model is related to the true size of the ICH, thus indicating differing performance by the volume of ICH. We see that most networks (except for 32) and PItcHPERFeCT perform similarly for ICH volumes up until about 100mL, where ICH volume > 100mL, the model does underestimate the true volume. We see that the 512 network does not suffer from this underestimation; the 3D nature of the other networks may be the cause.

**Table 3:** Volume statistics of automated and manual ICH volumes on the MISTIE III dataset. For each model, comparing the estimated volume to the manual volume, we present the correlation, root mean squared error (RMSE), percentage of patients with an estimated volume within 5mL of the manual, and the average volume difference, where negative values indicate the manual volume was larger than the automated. We see high correlation for all models but the 32 network. We used a threshold of 5mL as this was a measure of both ICH stability and clinically meaningful bleeding (size change) in the MISTIE protocol (defined as a volume change from concurrent scans). We note that the 64 network performs best on most measures for volume comparison.

| Model | Volume Correlation | RMSE | Percent with 5mL | Average Volume Difference |
| --- | --- | --- | --- | --- |
| **2D VNET 512** | 0.97 | 5.7 | 74% | -2.3 |
| **3D VNET 128** | 0.98 | 5.7 | 72% | -3.5 |
| **3D VNET 64** | **0.98** | **4.7** | **80%** | **-1.7** |
| **3D VNET 32** | 0.86 | 11.2 | 48% | -3.1 |
| **PltchPERFeCT** | 0.94 | 8.4 | 56% | -4.0 |

Given the overall linear relationship between the manual and automatic estimation in the top-performing models, we believe that these estimates should be good automated surrogates in analysis.

## Discussion

The vast majority of studies that employ DNN frameworks for ICH on CT are designed to classify the images in order to improve the sensitivity of detecting hemorrhage rather than perform volumetric segmentation(Chilamkurthy et al. 2018). A small number of prior peer reviewed studies have been published that automate segmentation of ICH with DNNs. In recent conference proceedings Islam et al.(Islam et al. 2018) published their ‘ICHNET’ for volumetric hemorrhage segmentation with a customized 2D architecture with convolutional layers based on the VGG16 network. Their image preprocessing steps included brain extraction which we also found significantly improved our results during initial testing. They achieved an average Dice Coefficient of 0.88 on their single institutional dataset of patients with ICH alone. Chang et al.(Chang et al. 2018) published a customized 2D/3D mask R-CNN architecture that performed combined detection and segmentation trained on a very large population of 10,159 patients with a validation set of 682 patients from a single institution. They report an impressive Dice Coefficient of 0.93 for ICH alone and 0.86 for SDH. There is no mention of the presence of IVH on any scans in their training or testing set, which is unusual given that this occurred naturally in 41% of patients in our clinical trial data and is expected in any sizable population of ICH patients and is likely to occur with a frequency of 25-41%. As expected, the presence of IVH reduced the Dice Coefficients of our segmentations as it can be present in small quantities in gravity dependent areas of the ventricular system and may intermix with cerebrospinal fluid reducing the average density. Several automated ICH models include level-set thresholds that may not translate well to ICH patients with IVH(Prakash et al. 2012).

In regards to post-processing, Islam et al.(Islam et al. 2018) used a 3D fully connected Conditional Random Field (CRF) and a connected components threshold to remove small areas of false positive voxels and smooth the segmentation. In our case, we did not perform any post-processing steps in this analysis to show the performance of the DNN. There are multiple options for data post-processing such as smoothing, connected components analysis, additional models based on bagging or other boosting algorithms, dilation and erosion, and hole filling. These morphological or algorithmic approaches may give slight improvements but can also increase deployment time.

We showed in our results that narrowing the anatomic context presented to neural network has a significant effect on the performance of an ICH segmentation model. As hypothesized, the 32 network showed overestimation of ICH in the cortex and around areas of calcification in the ventricular system (Figure 3, Figure 4). We also saw that the 32 network had an underestimation of ICH when the internal texture had areas of low density, often seen in hyperacute hemorrhage. This may be due to difficulty distinguishing these low density areas from CSF filled spaces. There are several existing neural network architectures that utilize small initial patch sizes and selecting the right spatial window while keeping in mind the networks RF may help improve results. Importantly, we found that the non-DNN PItcHPERFeCT model, which includes both patch sampling and whole brain heuristics was able to outperform VNet when inadequate anatomic context was presented. We plan to continue to use PItcHPERFeCT for verification of automated segmentations and suggest that poorly selected DNN models could be underperforming compared to existing work if it is not selected and trained with sufficient domain knowledge. The main drawbacks to PItcHPERFeCT are pragmatic: the long run time and the large number of software dependencies, though the latter can be mitigated with container services such as Docker; the authors provided an example web application demonstrating this model (https://smart-stats-tools.org/shiny/ich_segment_all/).

**Figure 3:**
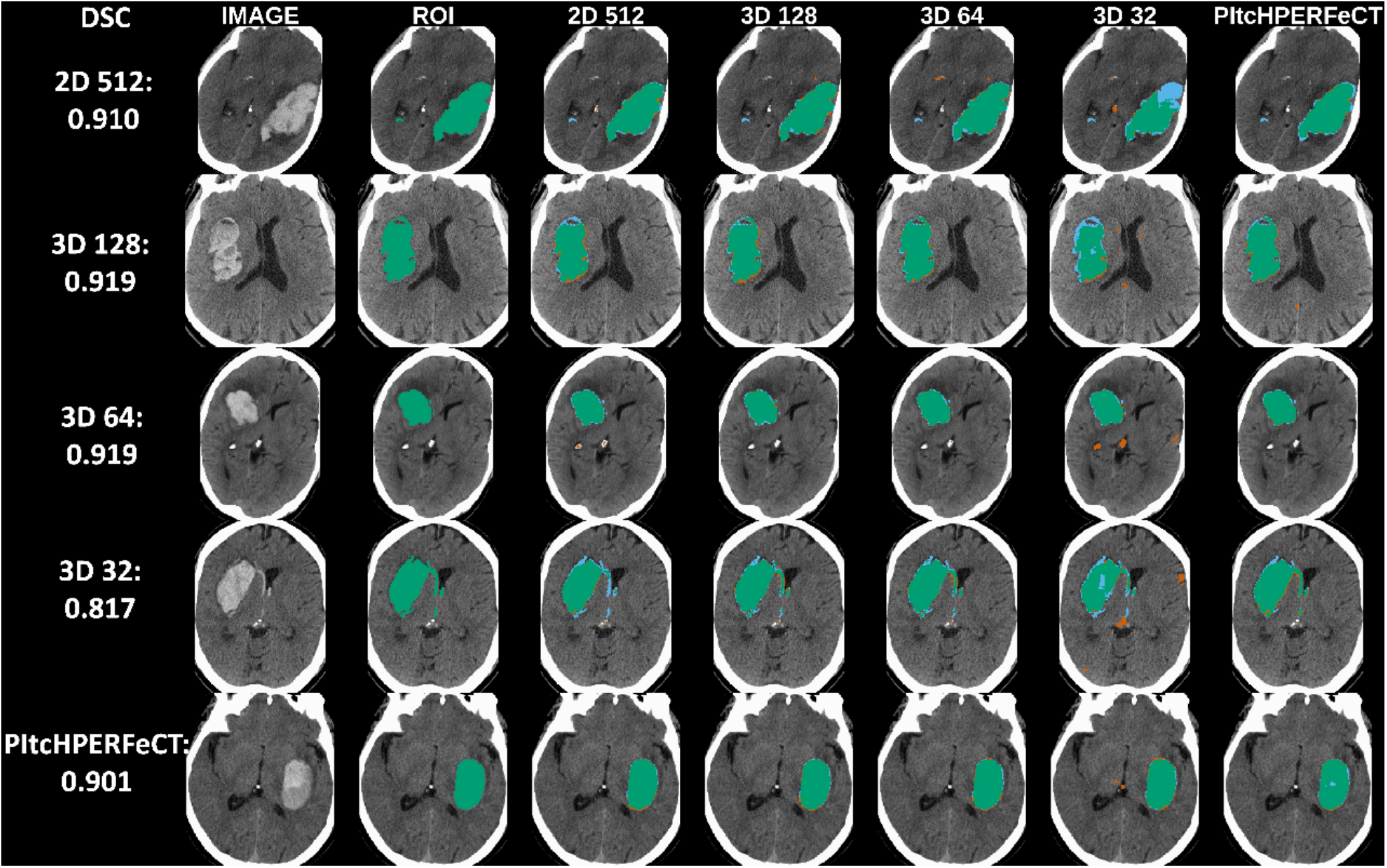
Median Segmentations and Images for Each Model. Each row represents the median segmentation for the network listed in the leftmost column (with the Dice coefficient for that case listed below). Areas colored in green represent a true positive, red represent a false positive and blue a false negative.

**Figure 4:**
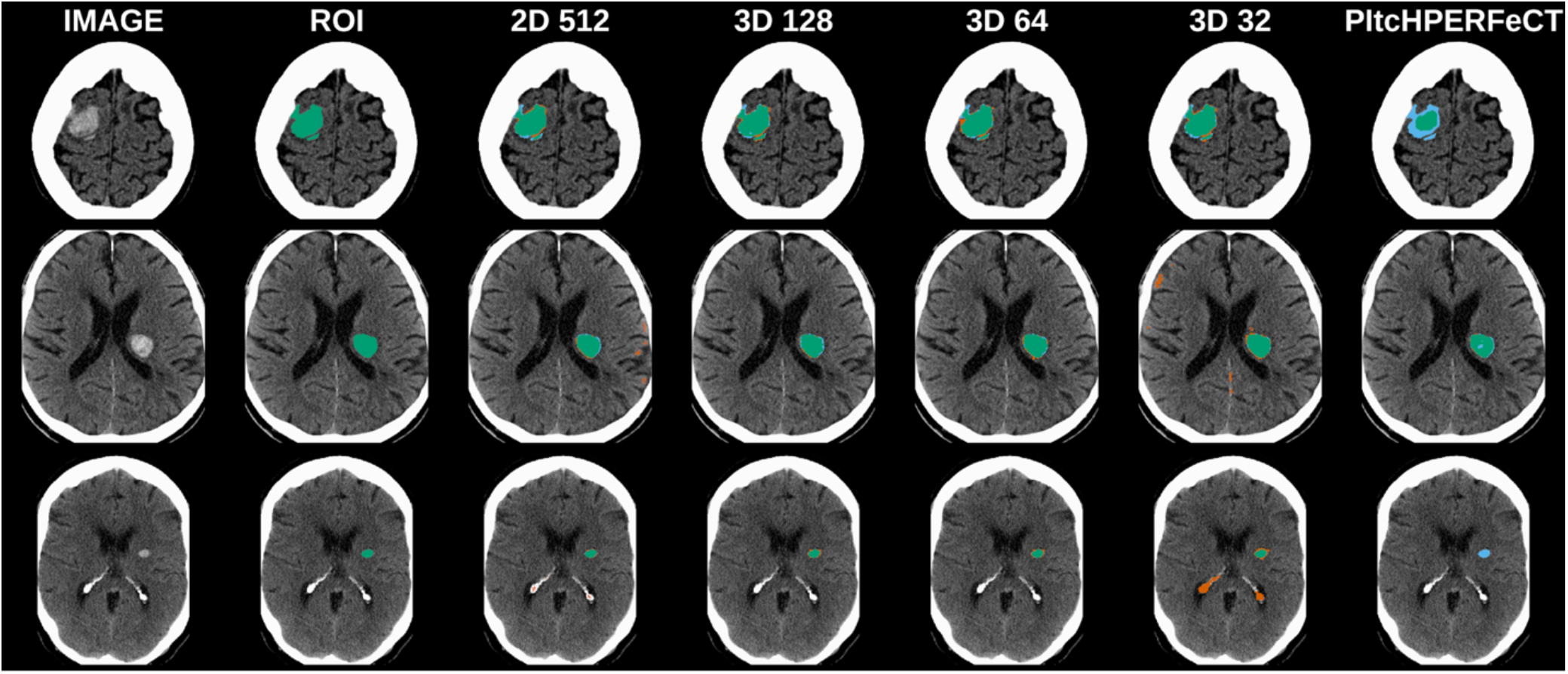
Examples of Failed Segmentation. Each row represents a different patient segmentation. Areas colored in green represent a true positive, red represent a false positive and blue a false negative. The first row represents a patient with a 12.3mL bleed, where PItchPERFeCT failed (DSC = 0.324), but the other methods performed adequately (DSC > 0.7). The second row is a patient with a 5.4mL bleed and was the patient where the 512 network failed with a DSC of 0.490. The last row shows a small ICH of 1mL, where PItchPERFeCT failed completely, but the deep networks segmented the blood, but the 32 network had a multiple false positives around calcifications in the ventricular system.

**Figure 5.**
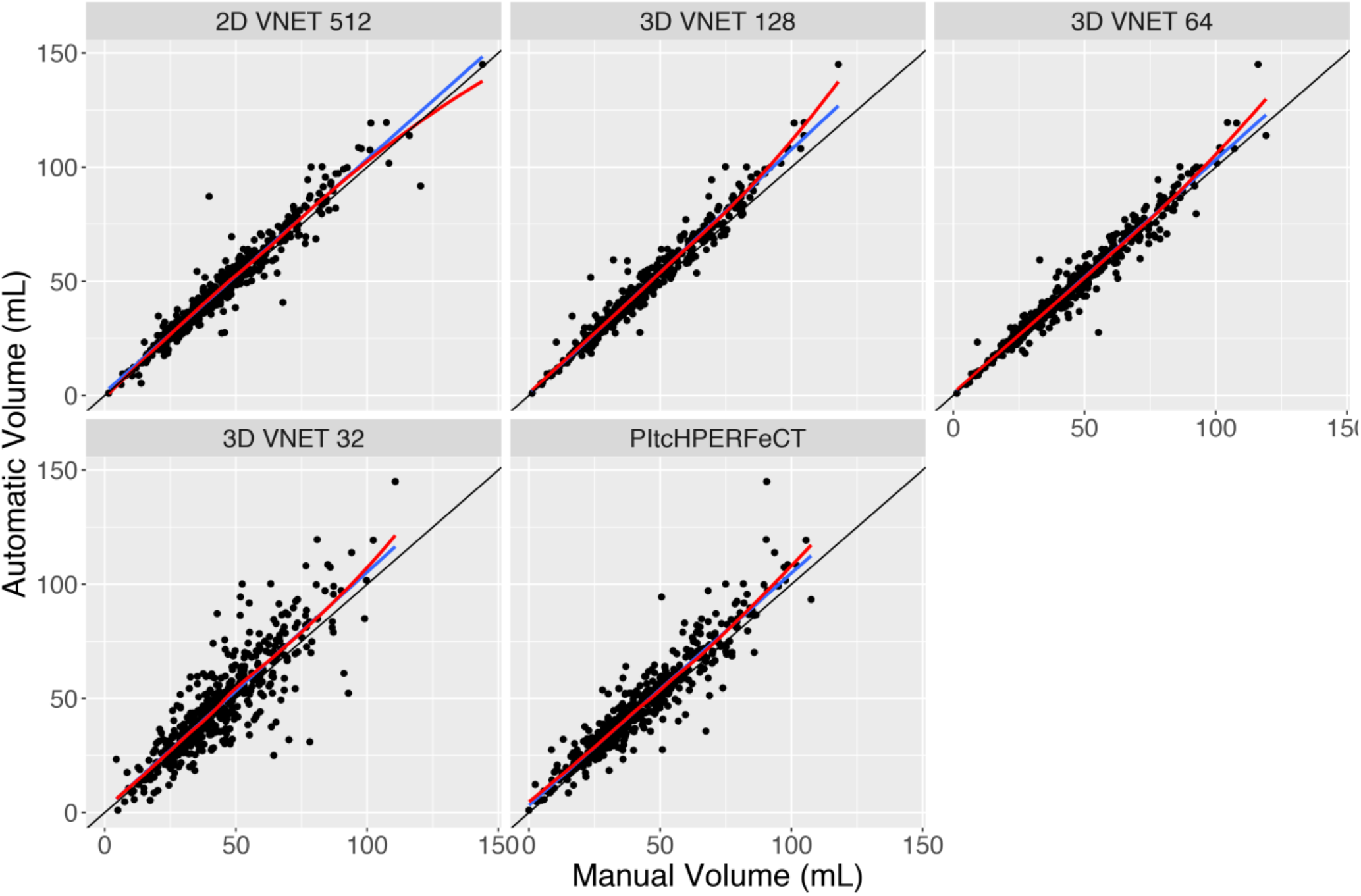
Comparison of manual and automated volumes from each model. Here we present the manual volume from gold standard hand segmentations from experts compared to the predicted volume from each automated model. The black line represents a X=Y line, the blue line represents a linear fit, and the red line represents a smoother (loess) fit. Here we see that the majority of the data falls close to the X=Y line, indicating a good correspondence between the automated and manual solutions. We also note the 3D VNET 32 has much more variability than the other VNET models.

**Figure 6.**
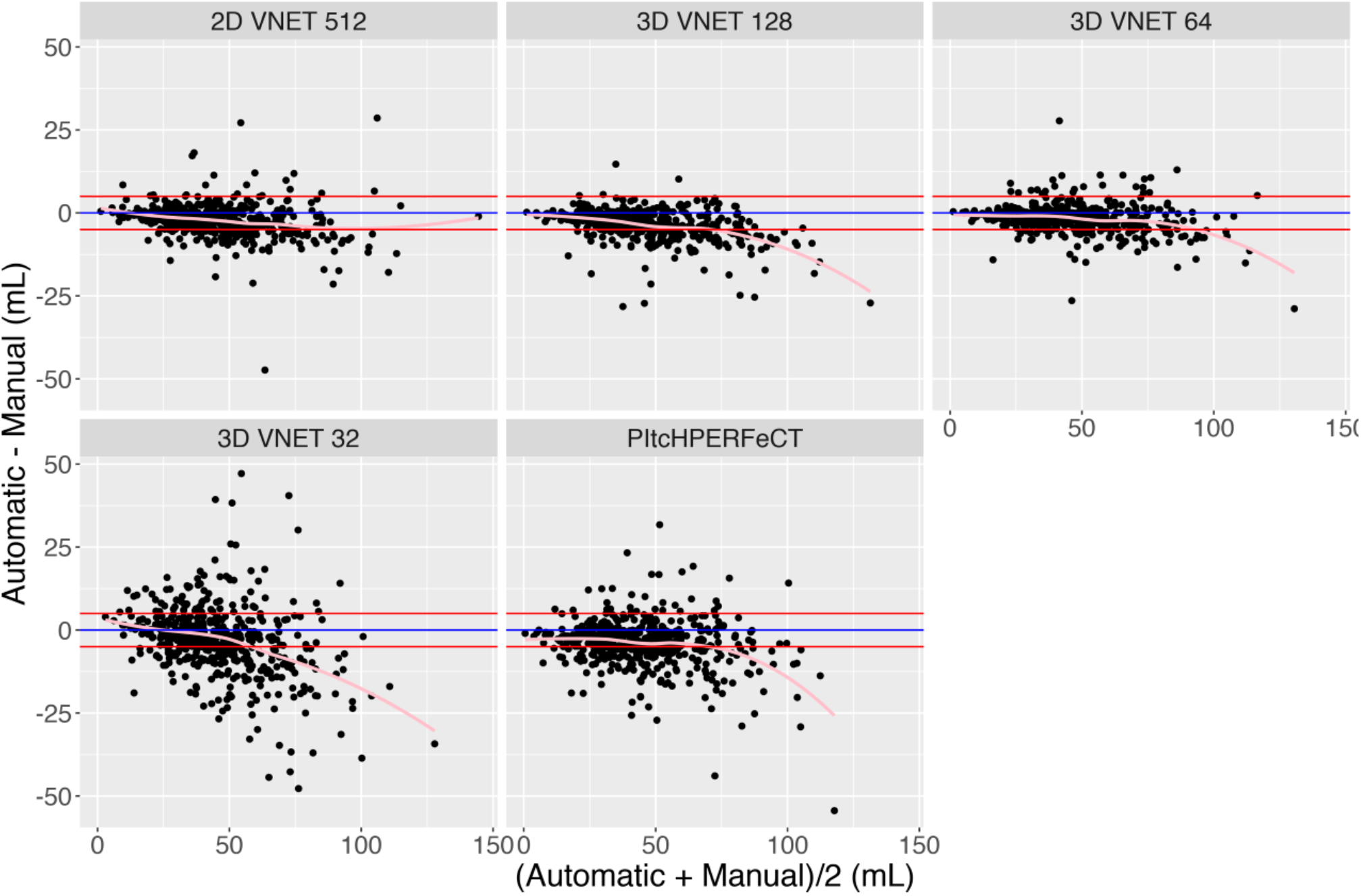
Bland-Altman plot of the automatic and manual segmentation volumes. The x-axis refers to the average volume of the automatic and manual segmentations and the y-axis is the difference, where negative values, refer to the automatic segmentation having a larger volume compared to the manual, gold standard. The blue line is no difference, the red lines are −5 and 5mL differences and the pink line is a smoother (loess) fit of the data. We see that the majority of the automatic volumes fall within 5mL of the manual volumes. The VNET 32 is not as accurate and the variability increases with volume. Though the PItcHPERFeCT method has reasonable accuracy, it is more variable and also does not perform as well. We see the 2D model and the 32 and 128 3D models perform reasonably for all volumes, though the 2D model does not have as much bias for very large volumes.

Another aim was to show that a DNN could be effectively trained to segment ICH in a smaller study as part of a clinical trial process. This required data augmentation and regularization techniques in order to avoid overfitting the network to the smaller data sample. Alternatively, we could have trained with the larger phase 3 study and validated on the smaller dataset or used cross validation, but we felt that curated and monitored data seen in a clinical trial progression of ICH would be more useful when considering application to new ICH therapeutic trials or other pathologies with brain hemorrhage, such as traumatic brain injury.

Limitations of this study are that the MISTIE clinical trials inclusion criteria were large, supratentorial ICH only. Thus the accuracy of the model for hemorrhages in the posterior fossa, where significant artifacts occur on CT is unknown. Another limitation is that epidural hemorrhage and subarachnoid w,ere not significantly observed in the dataset but could yield inaccurate results if co-occurring. Another limitation was that all segmentations were performed on noncontrast CT with radiation kernels designed for soft tissue. Sometimes in clinical research and practice, the only scan acquired or available is a non-contrast scan using a bone convolution kernel, contrasted CT or CT angiogram which we believe would yield significant false positive results.

In summary, we have shown that intracranial hemorrhage can be rapidly and accurately segmented from non-contrast CT scans utilizing a multi-center randomized control trial using either a 2D slice or 3D volumetric model. We performed a combination of standard neuroimaging procedures such as registration into template space and automated brain extraction and combined them with DNN techniques to optimize our networks. The 3D models in this paper could be easily trained registration-free like the 2D network. Resampling could also be done at a much higher resolution with higher GPU memory systems. Future directions for this research include further optimization through the process of pruning the network for resource efficient inference(Molchanov et al. 2016).

We believe our 2D 512 model has additional benefits over and above inference time. In practice, the entire brain may not be imaged or reconstructed from the PACS, but only a portion of the brain that is of interest with the hemorrhage. The 3D models may struggle with instances such as these due to the preprocessing steps of registration, as well as edge artifacts of the image. The 2D model does not suffer from these issues; but it relies on the 3D information in preprocessing steps such as skull-stripping.

Depending on the last layer of the model, one can estimate the probability of ICH at each voxel or a binary segmentation. Muschelli et al.(Muschelli et al. 2017) estimated the probability of ICH at the voxel level, determined a threshold to create a binary mask, and estimate ICH volume. In the CNN framework, we used sigmoid activations with a 1×1×1 convolution so that the result is binary, but estimating a better threshold from a probability, using a softmax layer, may improve performance in the future.

Our best results in terms of DSC and total volume calculated compared favorably with other published models trained on curated single center datasets but importantly include the neuroanatomic and neuropathological diversity and scanner acquisition variability often seen in ICH clinical practice. The natural history of ICH includes intraventricular extension of blood, particularly for hemorrhages close to the ventricles(Muschelli et al. 2015c) and the success of segmentation in this context has not previously been accounted for in segmentation studies based on either MRI or CT. This is a clear example of the need to understand the natural history of the underlying neuropathology as well account for the variability in acquisition when developing models for the clinical context, tasks that are frequently overlooked. This is especially so in the realm of DNNs where models with millions of parameters can be finely tuned to aspects of a curated dataset from a single institution that are not applicable externally. In our view, when decisions regarding potential therapeutic intervention are to be made, they should be informed by metrics and models validated in a prospective clinical trial on multicenter data designed with a full understanding of the underlying pathology.

## Data Availability

Publicly available software packages for immediate download for 3D Deep Neural Network Segmentation and
Preprocessing Python are hosted at https://github.com/msharrock/deepbleed and for the random forest model and
preprocessing in R at https://github.com/muschellij2/ichseg. The data archives for the MISTIE and MISTIE III
clinical trials are available through submission to the National Institute of Neurological Disorders and Stroke
Clinical Research Liaison https://www.ninds.nih.gov/Current-Research/Research-Funded-NINDS/ClinicalResearch/Archived-Clinical-Research-Datasets.

https://github.com/muschellij2/ichseg

## Supplemental

In this study, we trained end-to-end fully convolutional neural network strategies that have proven adept at handling medical imaging segmentation tasks. The UNet was originally developed to segment neuronal structures on 2D image slices and its 3D derivation and a derivative model, the VNet was developed to perform binary segmentation on isotropic 3D magnetic resonance images(Çiçek et al. 2016; Milletari et al. 2016). In each case, the left side of the network consists of a compression path and the right side a decompression path until the image reaches the input dimensions. The VNet consists of four compression stages, employing a progressive increase from one to three 5×5×5 convolutional filters and a parametric ReLU, each followed by 2×2×2 convolution of stride 2. The decompression path similarly consists of 2×2×2 deconvolutions of stride 2 followed by a progresive decrease from three to one 5×5×5 convolutional filters. The original model produced a two-channel prediction followed by a softmax filter, we instead utilized a 1×1×1 convolution with sigmoid activation. VNet and UNet both utilize skip connections from each downsampling layer directly to each upsampling layer to preserve larger scale volumetric information in the prediction. The 2D version of VNet is the same, but does not utilize the third dimension of the filters. All network structures were programmed in the Tensorflow Keras module and dataflow and model training were programmed in Tensorflow.

Preprocessing steps included conversion into the NIfTI format and brain extraction. We then ensured consistent image and label dimensions for feeding into each network by linear and nearest neighbor resampling respectively (Figure 1). CT scans provide density measurements in Hounsfield Units (HU) relative to water and do not require intensity normalization.

Our training strategy included the use of small initial learning rates and the adaptive moment estimation (Adam) stochastic optimizer as the networks converged quickly on the training data in the first 100 epochs, leading to poor performance on the testing data. In order to prevent overfitting, we utilized augmentation with random left-right flipping and random elastic deformation without z-axis deformations given the low z-axis resolution in our dataset. We used mini-batches to train the networks in all of the 2D and 3D implementations. Weighted sampling was used where the likelihood of sampling a voxel is proportional to the cumulative intensity histogram. We used the dice loss function as implemented in Milletari et al.(Milletari et al. 2016). We trained all networks for a minimum of 200 epochs and thereafter, a decision to stop training was made once the moving average loss on the testing data did not improve over the previous 10 epochs. The networks were trained using CPU multithreading and a Nvidia Tesla P100 GPU. The training time was approximately 16 hours to reach 200 epochs.

We chose the VNET architecture because it was designed for binary image segmentation. Other architectures such as UNET, DeepMedic, high-res 3DNET exist and may achieve similar results. We chose a batch size of 8 due to data size and efficiency; batches were randomly chosen.

Another choice was to do a 2D or 3D model. Some of the benefits of doing a 2D model is that each slice is segmented, similar to how experts perform the task, can be parallelized more efficiently, and can be done on data where the full scan is not available or is cut off at certain points. The 2D models do not take into account the 3D nature of the data, infer information from other slices, which experts do implicitly, and can be invariant to certain rotations.

## Information Sharing Statement

Publicly available software packages for immediate download for 3D Deep Neural Network Segmentation and Preprocessing Python are hosted at https://github.com/msharrock/deepbleed and for the random forest model and preprocessing in R at https://github.com/muschellij2/ichseg. The data archives for the MISTIE and MISTIE III clinical trials are available through submission to the National Institute of Neurological Disorders and Stroke Clinical Research Liaison https://www.ninds.nih.gov/Current-Research/Research-Funded-NINDS/Clinical-Research/Archived-Clinical-Research-Datasets.

